# Impaired sleep microarchitecture is associated with locus coeruleus degeneration in Parkinson’s disease

**DOI:** 10.1101/2025.04.06.25325309

**Authors:** Christopher E. J. Doppler, Nora Sembowski, Dean Plottka, Maximilian Hommelsen, Sinah Röttgen, Justus T. C. Schwabedal, Simon J. Schreiner, Gereon R. Fink, Per Borghammer, Stephan Bialonski, Michael Sommerauer

## Abstract

**Study objectives:** Sleep disorders are common non-motor symptoms of Parkinson’s disease (PD) that significantly impact patients’ quality of life. Specifically, alterations in sleep microstructure – such as reduced slow-wave activity and sleep spindles - are prevalent in PD. The locus coeruleus (LC), the brain’s primary source of noradrenaline, plays a pivotal role in regulating both sleep and wakefulness and is highly vulnerable to neurodegeneration in PD. This study explores whether disruptions in sleep microarchitecture in PD are linked to LC degeneration.

**Methods:** We assessed polysomnography for sleep macroarchitecture, EEG spectral power, and spindle density in 32 PD patients and 24 age- and sex-matched controls. In a subset of the sample, neuromelanin-sensitive MRI was performed, and LC neuromelanin contrast was correlated to sleep metrics.

**Results:** PD patients exhibited reduced slow-wave activity (*p* < 0.01), slow to fast frequency ratio (*p* < 0.01) and spindle density (*p* < 0.05) compared to HC subjects. LC neuromelanin contrast was diminished in PD patients (*p* < 0.05). Even though group differences were detected for slow-wave activity, a positive correlation between LC contrast and spindle density but not slow-wave activity was observed in the entire sample.

**Conclusions:** The findings indicate that spindle density, but not slow-wave activity, is associated with LC degeneration. Further research is needed to determine whether, besides this association, noradrenergic dysfunction is causal for impaired sleep microarchitecture and whether this connection also contributes to cognitive decline in PD and other neurodegenerative diseases, such as Alzheimer’s disease.

**Statement of significance:** This study provides novel evidence linking degeneration of the locus coeruleus with reduced sleep spindle density in Parkinson’s disease. While sleep macroarchitecture remains largely intact in Parkinson’s disease, our findings highlight the importance of investigating sleep microstructure to understand disease-related sleep disturbances. Given the role of the locus coeruleus in regulating sleep and cognition, our results suggest that sleep spindle alterations may serve as a biomarker for noradrenergic dysfunction. Future research should explore whether these changes contribute to cognitive decline in Parkinson’s disease and other neurodegenerative diseases. Longitudinal studies are needed to assess the potential of sleep microstructure as an early marker of disease progression and a target for therapeutic interventions.

## Introduction

Sleep disorders are common non-motor symptoms of Parkinson’s disease (PD) and have a substantial impact on patients’ quality of life. [1,2] Surprisingly, the disparities in macroparameters of sleep between PD patients and healthy aged volunteers are rather small. A meta-analysis combining 63 polysomnographic studies revealed a difference of less than 3 % in REM (rapid eye movement) and NREM (non-REM) sleep stages. [3] However, studies have shown that sleep spindle density [4–8] and slow-wave activity are reduced in PD [9,10] and are associated with cognitive decline in PD. [7,8,10,11] Hence, investigating sleep microstructure could facilitate a more comprehensive understanding of the pathophysiology underlying disrupted sleep and cognitive impairment in PD. [12–16]

There is mounting evidence of noradrenergic dysfunction in PD [17–20], which is also believed to contribute to cognitive decline in PD [17,21,22], a finding that has been well-documented in the context of Alzheimer’s disease. [23]. Neuropathological studies indicate that the locus coeruleus (LC), the brainstem structure that serves as the primary source of noradrenaline in the brain, is highly susceptible to PD pathology and is affected early in the course of the disease. [17,18,24,25] Furthermore, the LC has been identified as a key player in the regulation of sleep. [26,27] However, the role of the LC in alterations of microsleep in PD remains unclear. Given the impact of the LC on both cognition and sleep physiology, noradrenergic dysfunction may be the underlying mechanism for the influence of non-REM sleep disruption on progression in PD. Thus, the objective of our current study was to investigate whether changes in sleep spindle density and slow-wave activity were associated with LC degeneration.

## Materials and Methods

### Participants

Seventy-one datasets from two case-control studies, including deep phenotyping and polysomnography were pooled for analysis. [17,28,29] To avoid potential confounds due to comorbidities, we excluded datasets from 6 healthy control (HC) subjects and 9 PD patients due to high apnea/hypopnea indices exceeding age-matched reference values (AHI, >20/hour). [30] The final sample consisted of 24 HC subjects and 32 PD patients, all of whom were between 50 - 85 years old and non-demented (Montreal Cognitive Assessment score > 22). PD diagnosis was established following the current consensus criteria. [31] Levodopa equivalent daily doses (LEDD) were calculated as previously proposed. [32] Disease severity was judged according to the Hoehn and Yahr stage [33], and motor symptoms were quantified using the MDS Unified Parkinson’s disease Rating Scale part III (MDS-UPDRS III) after 12 hours of medication withdrawal.

The studies were approved by the local ethical committees, and all subjects provided informed written consent in accordance with the Declaration of Helsinki.

### Polysomnographic data and analysis of sleep microarchitecture

Overnight video-polysomnography (PSG) was conducted using a mobile SOMNOscreen™ plus device, including 10 EEG recordings (according to the international 10/20 system: F3, F4, C3, C4, O1, O2, M1, M2, Fpz as grounding, and Cz as reference) as previously described. [12,13] Visual PSG scoring was performed on 30-second epochs according to the AASM Manual for the Scoring of Sleep and Associated Events, Version 3. [34] We investigated sleep efficiency, sleep latency, absolute and relative time spent in each sleep stage, apnea-hypopnea indices, and periodic limb movement indices. Artifacts were marked manually.

### Spectral power

Polysomnographic recordings were exported as EDF (European Data format) files and analyzed using custom-made Python scripts. First, EEG data was preprocessed by applying a 0.5 Hz high-pass and 40 Hz low-pass filter and resampled to 128 Hz. Visually scored artifacts and arousals were then removed from the analysis. We performed a Fast Fourier Transformation of 5 seconds long segments to determine the spectral power of the delta (0.5 - 4 Hz), theta (4 – 8 Hz), alpha (8 - 12 Hz), sigma (12 – 15 Hz) and beta (15 - 32 Hz) frequency bands. Given that the objective of this analysis - based on our hypotheses and previous literature - was to examine slow-wave activity, further calculations were conducted using delta power exclusively. The extraction of delta power was performed using electrodes F3 and F4 (both referenced to the contralateral mastoid) during sleep stages N2 and N3 for spectral analyses, consistent with previous studies. [9] To avoid a potential bias due to interindividual differences in overall EEG amplitude, we normalized spectral power of the respective frequency band to total spectral power (0.5 – 32 Hz). [35]. The power from left and right hemisphere was averaged for further analysis. Additionally, slow-to-fast frequency ratios were calculated from absolute spectral power values ([delta]/[theta + alpha + beta + sigma]).

### Spindle analysis

Spindle detection was carried out using a validated deep neural network (SUMO) [36]. To prepare the polysomnographic recordings for automated detection, signals were first resampled at 100 Hz and filtered with an 8th-order, phase-preserving Butterworth filter (0.3– 30 Hz passband). Recordings were then normalized by z-transforming 115-second-long consecutive segments for each electrode to zero mean and unit variance, replicating the preprocessing used by SUMO [32]. SUMO detected sleep spindles from electrodes C3 and C4 (referenced to the contralateral mastoid), and only those occurring during sleep stage N2 were analyzed (detections in other stages, arousals, or artifact regions were discarded). Spindles lasting less than 0.5 s were discarded, following the minimum duration criterium for spindles according to the AASM guidelines [34]. To estimate the mean amplitude and frequency of each spindle, we followed a method from a prior study [37]. Spindle segments were bandpass filtered between 10–16 Hz using a zero-phase 4th-order Butterworth filter. The Hilbert transform was applied to compute instantaneous amplitudes, and the mean amplitude of a spindle was determined as the average of instantaneous amplitudes over the spindle duration. The frequency of a spindle was determined by identifying negative-to-positive zero-crossings of the filtered signal, calculating local frequencies as reciprocals of zero-crossing intervals, and averaging those local frequencies with associated instantaneous amplitudes above 3 µV to exclude likely non-spindle oscillations. Spindles without instantaneous amplitudes above 3 µV were discarded. For further analysis, parameters derived from electrodes C3 and C4 were averaged.

### Neuromelanin-sensitive MRI

We acquired 2D axial turbo spin-echo (TSE) T1-weighted sequences in a subset of 29 Parkinson’s disease patients and 13 HC subjects on a Siemens Trio 3T MR scanner using an 8- channel head coil with the following protocol: repetition time/echo time: 600 ms/9.2 ms, 26 averages, voxel size: 0.7 × 0.7 × 2 mm^3^). Planes were acquired perpendicular to the dorsal brainstem. Four images (one HC subject and three PD patients) had to be discarded due to artifacts.

Neuromelanin MRI contrast of the LC was quantified using a voxelwise analysis. TSE images were sinc-interpolated using PMOD 4.0 to achieve near-isotropic voxel dimensions of 0.5 × 0.7 × 0.7 mm^3^. For coregistration to a study-specific template space, individual TSE images were padded by 50 voxels in in all dimensions. A study-specific template was generated using the ‘antsMultivariateTemplateConstruction2’ function within Advanced Normalization Tools (ANTs v2.3.1). [38] Three expert raters manually segmented the LC on the resulting template (CEJD, SR, MS), identifying hyperintense voxels bilaterally within the dorsal pons contiguous to the fourth ventricle using ITK-SNAP. [39] A mask of the LC was created based on all voxels for which a consensus in manual segmentation could be achieved between at least two of the three raters. Voxel intensities of individual TSE images were normalized to the pons background VOI. Group differences in LC neuromelanin contrast as well as correlation between spindle density/slow-wave activity and LC neuromelanin contrast including all subjects were computed voxelwise using nonparametric permutation inference with FSL’s randomize, applying 5000 permutations after smoothing with a sigma of 2 mm. [40] Threshold-free cluster enhancement (TFCE) statistics were used, and FDR correction was applied to account for multiple comparisons with a threshold of *p* < 0.05. In addition to the voxelwise analysis of the data, following previous studies [17,41], we also computed the mean of the 10 voxels with the highest intensity inside the LC mask for each subject.

### Statistical analysis

We analyzed the data with RStudio 2024.12.0+467. Group data are presented as mean ± standard deviation unless otherwise stated. The normal distribution of the data was assessed using the Shapiro-Wilk test, Q-Q plots, and box plots. Group comparisons were performed using *t*-tests, Mann-Whitney-U-tests, and Pearson’s chi-squared tests as appropriate. Significance was accepted at *p* < 0.05.

## Results

### Demographic and clinical data

PD patients had an average age of 67.2 ± 7.1 years, and 28 % of them were female. The average disease duration was 7.4 ± 5.6 years, and Hoehn and Yahr stage was 2.4 ± 0.7. HC subjects did not differ significantly for age and sex (Table 1).

**Table 1:** Demographic and clinical characteristics and metrics of sleep macroarchitecture.

|  | HC<br>(n = 24) | PD<br>(n = 32) | p |
| --- | --- | --- | --- |
| Age [y] | 66.8 ± 7.3 | 67.2 ± 7.1 | 0.837 |
| Sex [f/m] | 8/16 | 9/23 | 0.900<sup>a</sup> |
| BMI [kg/m<sup>2</sup>] | 25.4 ± 2.6 | 25.8 ± 4.5 | 0.888<sup>b</sup> |
| *Clinical characteristics* |  |  |  |
| Time since diagnosis [y] |  | 7.4 ± 5.6 |  |
| MDS-UPDRS III (ON) |  | 29.6 ± 15.1 |  |
| MDS-UPDRS III (OFF) |  | 40.7 ± 16.7 |  |
| Hoehn & Yahr stage |  | 2.4 ± 0.7 |  |
| LEDD [mg] |  | 641.0 ± 469.5 |  |
| MoCA | 27.3 ± 1.6 | 26.7 ± 2.1 | 0.231<sup>b</sup> |
| *Sleep macroarchitecture* |  |  |  |
| Sleep efficiency [%] | 80.7 ± 12.8 | 79.4 ± 19.8 | 0.766<sup>b</sup> |
| Sleep latency [min] | 14.4 ± 14.7 | 19.2 ± 50.4 | 0.180<sup>b</sup> |
| Wake [min] | 90.6 ± 54.4 | 101.9 ± 81.5 | 0.941<sup>b</sup> |
| N1 [min] | 75.4 ± 38.4 | 58.9 ± 25.3 | 0.090<sup>b</sup> |
| N2 [min] | 181.5 ± 55.7 | 177.6 ± 68.6 | 0.812 |
| SWS [min] | 69.6 ± 27.9 | 56.0 ± 33.0 | 0.107 |
| REM [min] | 57.7 ± 24.0 | 63.6 ± 39.4 | 0.518 |
| AHI [/h] | 8.1 ± 6.8 | 6.7 ± 6.6 | 0.337<sup>b</sup> |
| PLMI | 24.0 ± 36.0 | 36.4 ± 38.5 | 0.138<sup>b</sup> |
| RBD [y/n] | 0/24 | 14/18 | <0.0001<sup>a</sup> |

**Abbreviations:** AHI = apnea-hypopnea index, BMI = body mass index, f = female, h = hour, HC = healthy controls, kg = kilogram, LEDD = levodopa equivalent dose, m = male or meter, MDS-UPDRS III = Movement Disorders Society Unified Parkinson's disease Rating Scale part III, mg = milligram, min = minutes, N = non rapid eye movement sleep, PD = Parkinson's disease patients, PLMI = periodic limb movement index, RBD = REM sleep behavior disorder, REM = rapid eye movement sleep, s = second, SWS = slow wave sleep, y = years; <sup>a</sup>Pearson's chi-squared test, <sup>b</sup>Mann-Whitney-U-test.

We did not observe statistically significant differences in sleep macroarchitecture parameters between HC subjects and PD patients, including sleep efficiency, sleep latency, sleep stages, AHI, and PLMI (Table 1).

### Analysis of sleep microstructure

In PD patients, spindle density was significantly reduced (0.6 ± 0.5 versus 1.1 ± 1.1, *p* = 0.021, Table 2, Figure 1C). There we no differences in spindle configuration, including amplitude and duration, between the two groups.

**Table 2:**
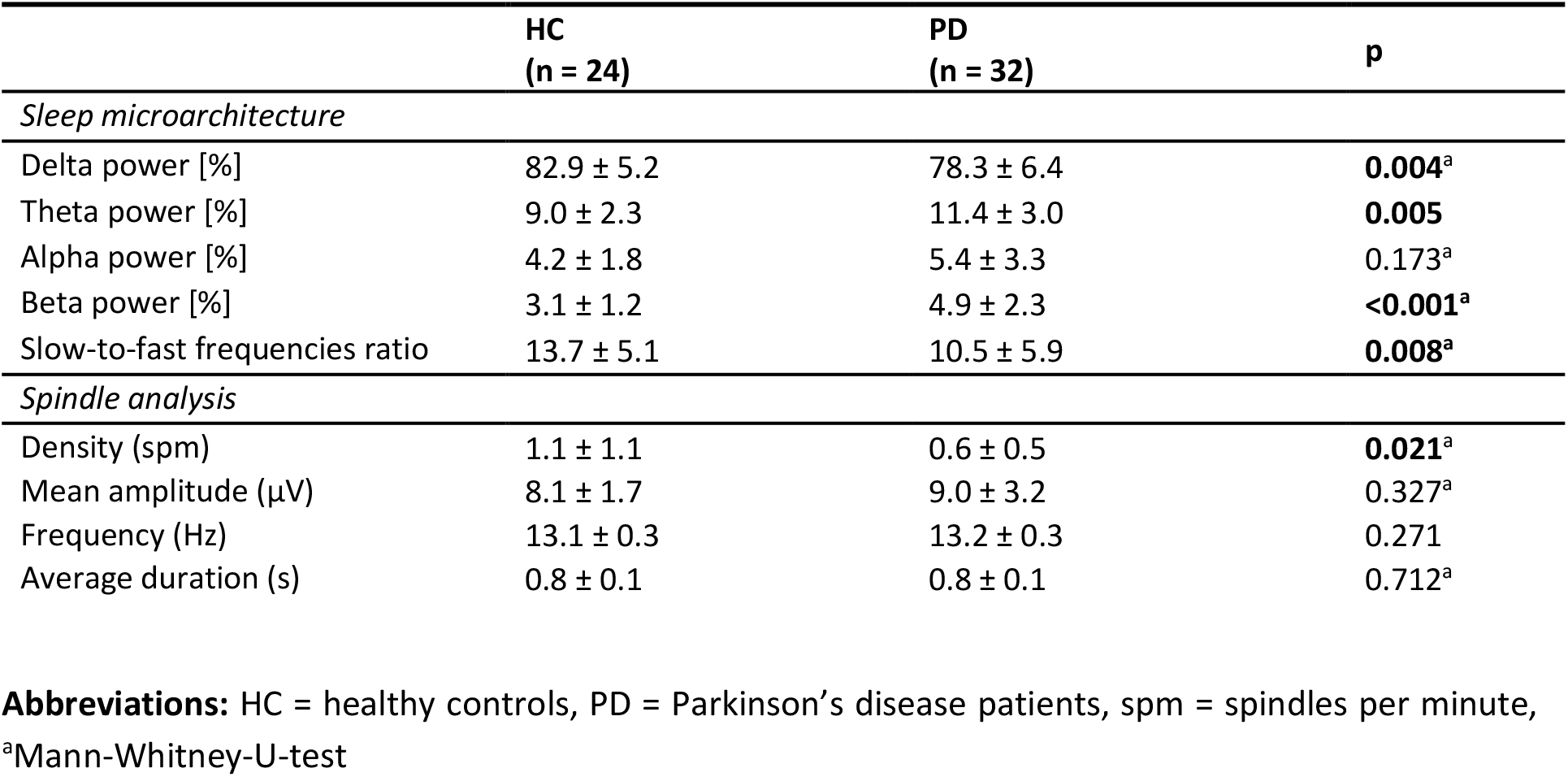
Metrics of sleep microarchitecture.

**Figure 1:**
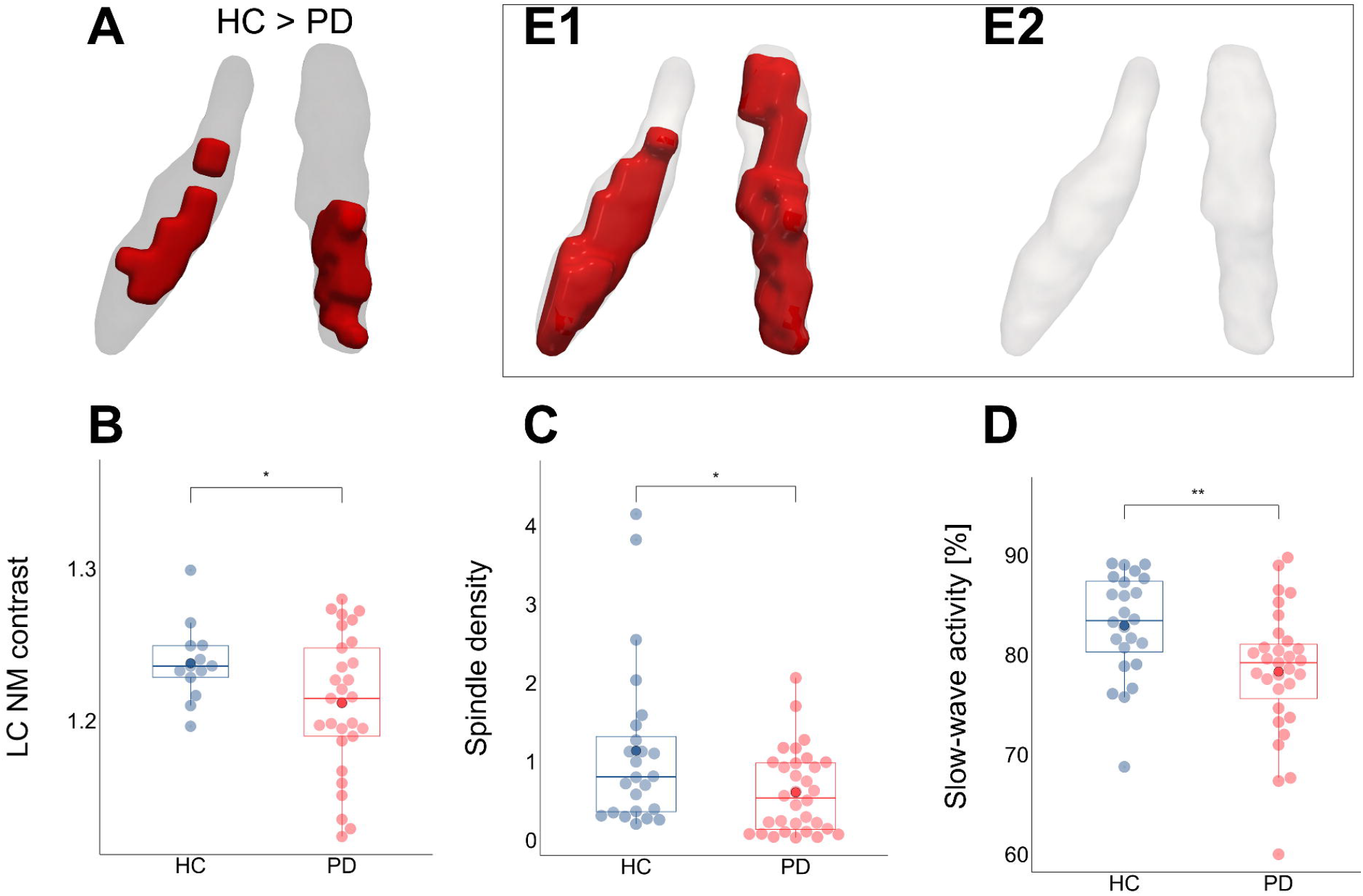
**Group differences between Parkinson’s disease (PD) patients and healthy controls (HC) in locus coeruleus (LC) neuromelanin (NM) contrast (as derived from voxelwise analysis (A) and extraction of the mean value of the 10 voxels with the highest intensities (B)), spindle density (C), and slow-wave activity (D). Inset: voxelwise correlations between spindle density (E1) as well as slow-wave activity (E2) and LC neuromelanin contrast**. The LC mask is depicted in dark grey for group comparisons and light grey for correlations, significant voxels (according to threshold-free cluster enhancement (TFCE) statistics with 5000 permutations and after FDR correction) are shown in red (A, E1, E2). For illustrative purposes both statistical maps were resampled to a voxel size of 0.25 mm^3^ isotropic and binarized. The left LC is shown on the left side of the image. Slow-wave activity (derived from EEG electrodes F3/F4 and normalized to total spectral power, D) and spindle density (derived from EEG electrodes C3/C4 during sleep stage N2) are reduced in PD patients (red dots) compared to HC (blue dots). Mean values are depicted as singular dots with higher saturation. (* *p* < 0.05, ** *p* < 0.01).

PD patients exhibited reduced slow-wave activity during consolidated sleep compared to HC subjects (78.3 ± 6.4 versus 82.9 ± 5.2 %, *p* = 0.004, Table 2, Figure 1D). Consequently, fractions of the remaining frequency bands, besides the alpha band, were increased (theta, *p* < 0.01; beta, *p* < 0.001), resulting in a lowered slow to fast ratio in PD patients (10.5 ± 5.9 versus 13.7 ± 5.1, *p* = 0.008). Comparable findings were obtained when analyzing sleep stages N2 and N3 separately (Supplementary Table 1).

### Locus coeruleus neuromelanin contrast

According to both the voxelwise analysis (Figure 1A) and the extraction of the 10 voxels with the highest intensities (Figure 1B), PD patients exhibited a decrease in LC neuromelanin contrast compared to HC subjects.

Furthermore, a positive voxelwise correlation between spindle density and LC neuromelanin contrast was observed across all subjects in large parts of the right LC and middle and caudal portions of the left LC (Figure 1E1). However, we did not observe a correlation between LC neuromelanin contrast and slow-wave activity (Figure 1E2).

## Discussion

Our findings revealed a decrease in slow-wave activity and spindle density in PD patients compared to HC subjects. In contrast, sleep macroparameters did not differ between the two groups. Additionally, we could link noradrenergic deterioration specifically with reduced spindle density. However, even though slow-wave activity was reduced in PD patients, this parameter was not associated with LC neuromelanin contrast. This suggests that spindle density depends on integrity of the locus coeruleus.

Changes in sleep microstructure, specifically a decrease in slow-wave activity and spindle density, are part of physiological aging. [42] However, in neurodegenerative disorders such as PD, these changes are significantly amplified and associated not only with prevalent subjective sleep disturbances but also cognitive complaints. [4–7,9,10,43] The link between disrupted sleep and neurodegeneration is also considered to be bidirectional. [44] For PD, an association between disease progression and reduced slow-wave activity has been shown. [45]. In animal studies, slow-wave activity correlated with glymphatic influx, [46] suggesting a mechanism by which sleep disturbances, through reduced clearance of toxic compounds such as pathologic α-synuclein species in PD, might promote disease progression. [47] This data corroborates the use of microsleep parameters as potential biomarkers for accelerated disease progression in PD.

The noradrenergic system also plays a crucial role in regulating non-REM sleep [26,27] and glymphatic clearance. [48] During sleep, activation of the LC is significantly reduced [49–51], and it serves to mediate awakening upon sensory stimuli. [52,53] However, even during non-REM sleep, a tonic firing pattern of the LC persists [49]. Changes in LC activity are important for the formation of sleep spindles in rats. [54] Additionally, in a mouse model, these changes are involved in infraslow oscillations, which alternate between periods of consolidated sleep (rich in sleep spindles) and vigilant sleep. [55] The integrity of the LC is a determining factor in sleep microarchitecture [56–58] and may therefore be considered the underlying pathomechanism for the observed reduction in spindle density in our sample. For cognition the noradrenergic system is considered to be a key player [21] and consequently degeneration of the LC, the major source of noradrenaline in the brain, is associated with cognitive deficits in PD and Alzheimer’s disease [59–61].

To the best of our knowledge, we provide the first evidence of an association between LC degeneration and reduced sleep spindle density in humans. Taken together with the evidence on the link between noradrenergic dysfunction and cognitive deficits and data from animal studies [62], it can be hypothesized that the LC might be the mediator between microsleep changes and cognition.

The cross-sectional nature of our data represents a limitation of our study. Longitudinal data on changes in microsleep parameters and in-depth neuropsychological testing during disease progression could provide further evidence for the association with noradrenergic dysfunction.

In spindle detection, manual scoring is considered the gold standard. However, this approach is highly time-consuming and only yields moderate inter-rater variability. [63] Automatic spindle scoring systems are available, but mostly rely on detecting specific physical features of spindles, such as amplitude and frequency. [64] In recent years, artificial intelligence techniques using neural networks have been shown to perform better than feature-based methods for event detection. [65] In this study, we used a recently proposed advanced spindle detection algorithm, SUMO (Slim U-Net trained on MODA), which is based on a deep neural network model. [36] SUMO outperformed available conventional algorithms and the majority of experts in the largest available manually scored dataset on spindles, MODA (Massive Online Data Annotation). [66] Thus, our data suggest that automatic spindle detection based on deep learning is feasible even in PD patients and may be superior to conventional algorithms.

In conclusion, we observed an association between microsleep parameters and degeneration of the locus coeruleus in PD patients. Further studies are warranted to investigate if noradrenergic dysfunction is causal for impaired sleep microarchitecture and whether this association is also related to cognitive decline in PD and other neurodegenerative diseases, such as Alzheimer’s disease.

## Acknowledgments

C. E. J. Doppler received grants from the Clinician Scientist Program (CCSP), funded by the German Research Foundation (DFG, FI 773/15-1).

M. Sommerauer received grants from the Else Kröner-Fresenius-Stiftung (grant number 2019_EKES.02), and funding from the program “Netzwerke 2021”, an initiative of the Ministry of Culture and Science of the State of North Rhine Westphalia. The Federal Ministry of Education and Research (BMBF) is funding the project within the framework of the funding programme ACCENT (funding code 01EO2107).

G.R.F. is funded by the Deutsche Forschungsgemeinschaft (DFG, German Research Foundation) – Project-ID 431549029 – SFB 1451.

## Disclosure Statement

G. R. Fink serves as an editorial board member of Cortex, Neurological Research and Practice, NeuroImage: Clinical, Zeitschrift für Neuropsychologie, and DGNeurologie; receives royalties from the publication of the books Funktionelle MRT in Psychiatrie und Neurologie, Neurologische Differentialdiagnose, and SOP Neurologie; received honoraria for speaking engagements from Bayer, Desitin, Ergo DKV, Forum für medizinische Fortbildung FomF GmbH, GSK, Medica Academy Messe Düsseldorf, Medicbrain Healthcare, Novartis, Pfizer, and Sportärztebund NRW.

The other authors have no conflict of interest to report.

## Data availability

The data underlying this article will be shared on reasonable request to the corresponding author.

## Notes

### Author Declarations

Ethics committee of Aarhus University and University of Cologne gave ethical approval for this work.

