## Supplementary Table 1 for "Impaired sleep microarchitecture is associated with locus coeruleus degeneration in Parkinson’s disease"

### Supplement

#### Supplementary Table 1: Normalized slow-wave activity for sleep stages N2/N3 separately

| | **HC (n = 24)** | **PD (n = 32)** | **p** |
|---|---|---|---|
| Slow-wave activity N2 [%] | 77.0 ± 6.9 | 73.2 ± 6.9 | **0.047** |
| Slow-wave activity N3 [%] | 88.7 ± 3.9 | 86.6 ± 3.5 | **0.040** |

Abbreviations: HC = healthy control, N2 = non rapid eye movement sleep stage 2, N3 = non rapid eye movement sleep stage 3, PD = Parkinson’s disease
